# Succumbing to the COVID-19 Pandemic – Healthcare Workers not Satisfied and Intend to Leave Their Jobs

**DOI:** 10.1101/2020.05.22.20110809

**Authors:** Stephen X. Zhang, Jiyao Chen, Asghar Afshar Jahanshahi, Aldo Alvarez-Risco, Huiyang Dai, Jizhen Li, Ross Mary Patty-Tito

## Abstract

**Background:** Healthcare workers are under such a tremendous amount of pressure during the COVID-19 pandemic that many have become concerned about their jobs and even intend to leave them. It is paramount for healthcare workers to feel satisfied with their jobs and lives during a pandemic.

**Methods:** Between 10 to 30 April, 2020, 240 healthcare workers in Bolivia completed a cross-sectional online survey, which assessed their job satisfaction, life satisfaction, and turnover intention in the ongoing COVID-19 pandemic.

**Results:** The results revealed that their number of office days predicted job satisfaction, life satisfaction, and turnover intention, but the relationships varied by their age. For example, healthcare workers’ office days negatively predicted job satisfaction for the young (e.g. at 25 years old: b=-0.21; 95% CI: −0.36 to −0.60) but positively predicted job satisfaction for the old (e.g. at 65 years old: b=0.25; 95% CI: 0.06 to 0.44).

**Conclusions:** These findings provide evidence to enable healthcare organizations to identify staff concerned about job satisfaction, life satisfaction, and turnover intention to enable early actions so that these staff can remain motivated to fight the prolonged COVID-19 pandemic.

## 1. Introduction

In the COVID-19 pandemic, healthcare workers are experiencing unprecedented pressure from stressors including but not limited to enormous workload, virus exposure, inadequate personal protective equipment (PPE), moral dilemmas, work incivility, despair, isolation from family, and discrimination (Lai et al., 2020; Zhang et al., in press). An emergency physician in Washington DC, Thomas Kirsch, asked on March 24, 2020, “How much risk do healthcare workers have to take? Or, more bluntly: How many of us will die before we start to walk away from our jobs?” (Kirsch, 2020). Many healthcare workers stopped showing up at work once COVID-19 hit the US, due to the shortage of PPE (Cloherty, 2020). Such issues can be more severe in less developed economies. For instance, media reported over 100 nurses stopped coming to work on one single day in a hospital in South America (Galdos, 2020).

In this critical time of the COVID pandemic, it is paramount for healthcare workers to feel satisfied with their current jobs and lives without intention to leave their jobs. However, little research has studied the job-related variables of healthcare workers during the COVID-19 pandemic. The existing research on healthcare workers under COVID-19 has focused on workers’ mental health and well-being (c.f. a meta-analysis by Pappa *et al*. (in press)). This study presents the first attempt to document healthcare workers’ job satisfaction, life satisfaction, and turnover intention, and their predictors during the COVID-19 pandemic.

We predict the job-related outcome variables by not only the risk factors of mental health issues identified by the literature, but also healthcare workers’ job-related characteristics, such as office days, and whether they are temporary staff or redeployed during the COVID-19 pandemic. These predictors can help healthcare organizations to be more specific in developing evidence-based screening for mental health, job satisfaction, and turnover issues of their staff during the COVID-19 pandemic (Yang *et al*., 2020).

## 2. Methods

Bolivia, with the lowest GDP per capita in South America, lacks adequate health infrastructure (Arigho-Stiles, 2020). COVID-19 cases appeared in Bolivia on March 10, 2020, and on March 25, 2020, Bolivia declared COVID-19 a national health emergency. COVID-19 has since spread quickly, generating enormous pressure on the country’s healthcare workers. Media reported healthcare workers in several hospitals turned away COVID-19 patients out of concern over the limited resources in overcrowded facilities (Laing and Ramos, 2020). On April 23, 2020, Bolivian health workers protested publicly about shortages of PPE and other supplies, including body bags for the dead (Requena, 2020).

This study followed the American Association for Public Opinion Research (AAPOR) guideline. All participants agreed with their informed consent to enroll in the online survey. Strict confidentiality was assured in the data collection processes was voluntary. The study was approved by the ethics committee of Tsinghua University (20200322). Using a stratified sampling, we sent out our survey to 402 healthcare workers in 83 healthcare facilities (40 in La Paz, 11 in Santa Cruz, and 32 from other administrative regions) in Bolivia from April 10 to April 30, 2020, one month into the COVID-19 emergency in Bolivia. The survey was open to healthcare workers in healthcare institutions such as hospitals, clinics, first emergency responders, medical wards, nursing homes, dental clinics, and pharmacies. 240 health workers filled the entire survey, resulting in a response rate of 59.7%.

The healthcare workers reported their demographic characteristics such as age, gender, number of children, education level, daily exercise hours, and number of office days in the past week. We also asked whether they were temporary staff or were redeployed from their usual positions (i.e. from non-ICU to ICU) to deal with the COVID-19 pandemic. The outcome variables included healthcare workers’ job satisfaction, life satisfaction, and turnover intention.

*Job satisfaction*. We measured healthcare workers’ job satisfaction by an five-item scale (Brayfield and Rothe, 1951, Montazeri *et al*., 2011). These five items were scored on a 1 (strongly disagree) to 7 (strongly agree) Likert scale. Sample items include “I find real enjoyment in my work” and “most days I am enthusiastic about my work.” The Cronbach’s alpha was 0.80.

*Life satisfaction*. The respondents reported using the five-item scale of Satisfaction with Life (SWL) (Diener *et al*., 1985). These five items were scored on a 1 (strongly disagree) to 7 (strongly agree) Likert scale. Sample items include “in most ways my life is close to my ideal” and “the conditions of my life are excellent.” The Cronbach’s alpha was 0.81.

*Turnover intention*. We used the 5-point scale to measure healthcare workers’ turnover intention (Cammann *et al*., 1979). The three items were scored on a 1 (strongly disagree) to 5 (strongly agree) Likert scale. Sample items include “I often think about quitting my job with my present organization” and “it is very possible that I will look for a new job next year.” The Cronbach’s alpha was 0.85.

We used Stata 16.0 to summarize the variables and to identify the predictors of job satisfaction, life satisfaction, and turnover intention with a 95% confidence level.

## 3. Results

### 3.1 Descriptive findings

Table 1 shows that 72.9% of the 240 healthcare workers were female, and 8.3% were younger than 29 years old, 29.2% were between 30 to 39 years, 57.5% were between 40 to 59 years, and 5.0% were 60 years or older. Most participants (91.7%) had an undergraduate degree or higher. Less than a third (30.8%) were temporary staff, and 10.8% had been redeployed to deal with the COVID-19 crisis. Finally, 6.7% of participants worked in healthcare facilities seven days in the past week, 4.2% worked six days, 14.2% worked five days, and 71.3% worked between one to four days, and 3.8% worked zero days.

**Table 1.**
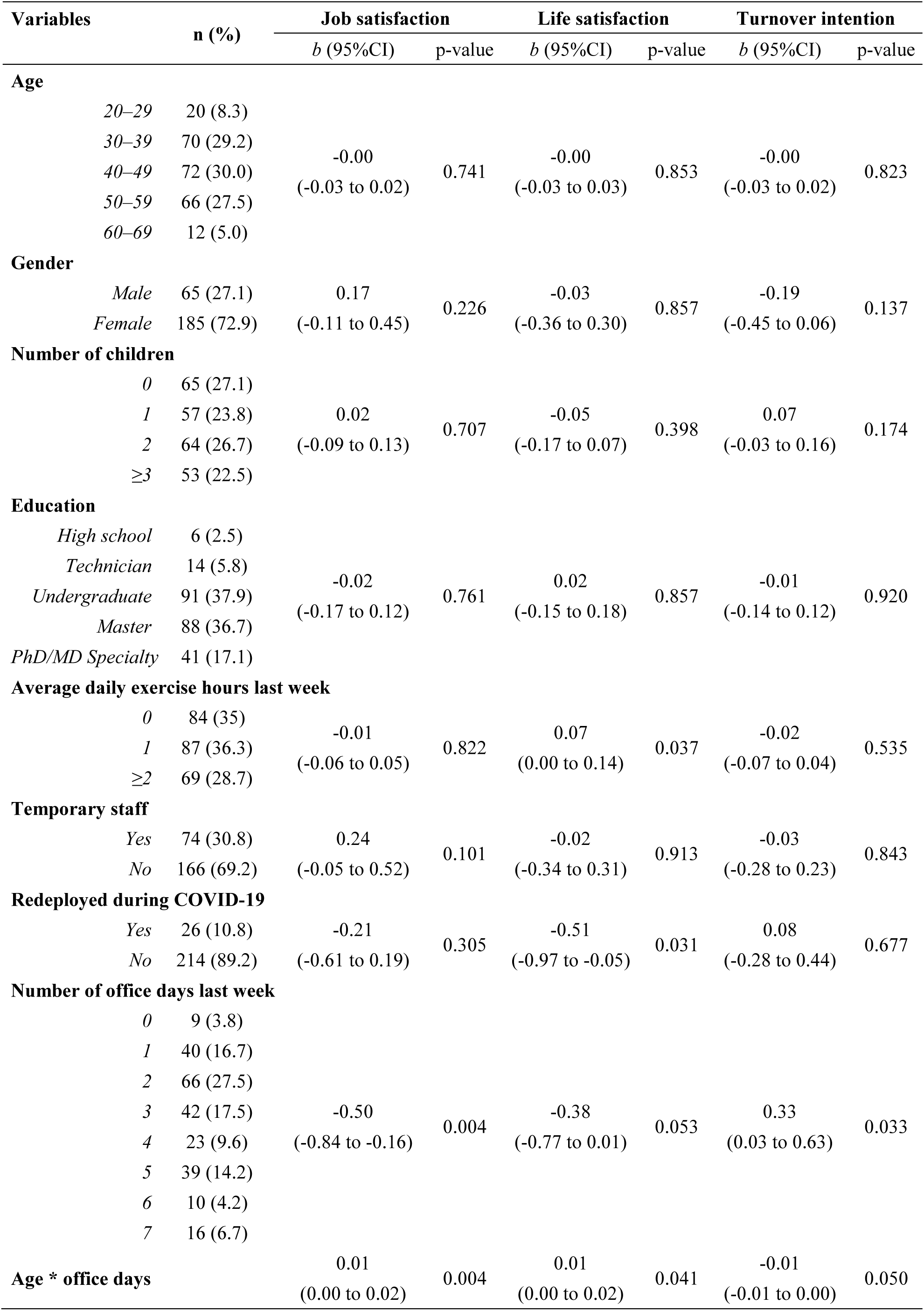
Predicting healthcare workers’ job satisfaction, life satisfaction, and turnover intention (N=240)

| Variables | n (%) | Job satisfaction |  | Life satisfaction |  | Turnover intention |  |
| --- | --- | --- | --- | --- | --- | --- | --- |
|  |  | b (95%CI) | p-value | b (95%CI) | p-value | b (95%CI) | p-value |
| Age |  |  |  |  |  |  |  |
| 20–29 | 20 (8.3) |  |  |  |  |  |  |
| 30–39 | 70 (29.2) |  |  |  |  |  |  |
| 40–49 | 72 (30.0) | -0.00 | 0.741 | -0.00 | 0.853 | -0.00 | 0.823 |
| 50–59 | 66 (27.5) | (-0.03 to 0.02) |  | (-0.03 to 0.03) |  | (-0.03 to 0.02) |  |
| 60–69 | 12 (5.0) |  |  |  |  |  |  |
| Gender |  |  |  |  |  |  |  |
| Male | 65 (27.1) | 0.17 | 0.226 | -0.03 | 0.857 | -0.19 | 0.137 |
| Female | 185 (72.9) | (-0.11 to 0.45) |  | (-0.36 to 0.30) |  | (-0.45 to 0.06) |  |
| Number of children |  |  |  |  |  |  |  |
| 0 | 65 (27.1) |  |  |  |  |  |  |
| 1 | 57 (23.8) | 0.02 | 0.707 | -0.05 | 0.398 | 0.07 | 0.174 |
| 2 | 64 (26.7) | (-0.09 to 0.13) |  | (-0.17 to 0.07) |  | (-0.03 to 0.16) |  |
| ≥3 | 53 (22.5) |  |  |  |  |  |  |
| Education |  |  |  |  |  |  |  |
| High school | 6 (2.5) |  |  |  |  |  |  |
| Technician | 14 (5.8) |  |  |  |  |  |  |
| Undergraduate | 91 (37.9) | -0.02 | 0.761 | 0.02 | 0.857 | -0.01 | 0.920 |
| Master | 88 (36.7) | (-0.17 to 0.12) |  | (-0.15 to 0.18) |  | (-0.14 to 0.12) |  |
| PhD/MD Specialty | 41 (17.1) |  |  |  |  |  |  |
| Average daily exercise hours last week |  |  |  |  |  |  |  |
| 0 | 84 (35) |  |  |  |  |  |  |
| 1 | 87 (36.3) | -0.01 | 0.822 | 0.07 | 0.037 | -0.02 | 0.535 |
| ≥2 | 69 (28.7) | (-0.06 to 0.05) |  | (0.00 to 0.14) |  | (-0.07 to 0.04) |  |
| Temporary staff |  |  |  |  |  |  |  |
| Yes | 74 (30.8) | 0.24 | 0.101 | -0.02 | 0.913 | -0.03 | 0.843 |
| No | 166 (69.2) | (-0.05 to 0.52) |  | (-0.34 to 0.31) |  | (-0.28 to 0.23) |  |
| Redeployed during COVID-19 |  |  |  |  |  |  |  |
| Yes | 26 (10.8) | -0.21 | 0.305 | -0.51 | 0.031 | 0.08 | 0.677 |
| No | 214 (89.2) | (-0.61 to 0.19) |  | (-0.97 to -0.05) |  | (-0.28 to 0.44) |  |
| Number of office days last week |  |  |  |  |  |  |  |
| 0 | 9 (3.8) |  |  |  |  |  |  |
| 1 | 40 (16.7) |  |  |  |  |  |  |
| 2 | 66 (27.5) |  |  |  |  |  |  |
| 3 | 42 (17.5) | -0.50 | 0.004 | -0.38 | 0.053 | 0.33 | 0.033 |
| 4 | 23 (9.6) | (-0.84 to -0.16) |  | (-0.77 to 0.01) |  | (0.03 to 0.63) |  |
| 5 | 39 (14.2) |  |  |  |  |  |  |
| 6 | 10 (4.2) |  |  |  |  |  |  |
| 7 | 16 (6.7) |  |  |  |  |  |  |
| Age * office days |  |  |  |  |  |  |  |
|  |  | 0.01 | 0.004 | 0.01 | 0.041 | -0.01 | 0.050 |
|  |  | (0.00 to 0.02) |  | (0.00 to 0.02) |  | (-0.01 to 0.00) |  |

**Figure 1.**
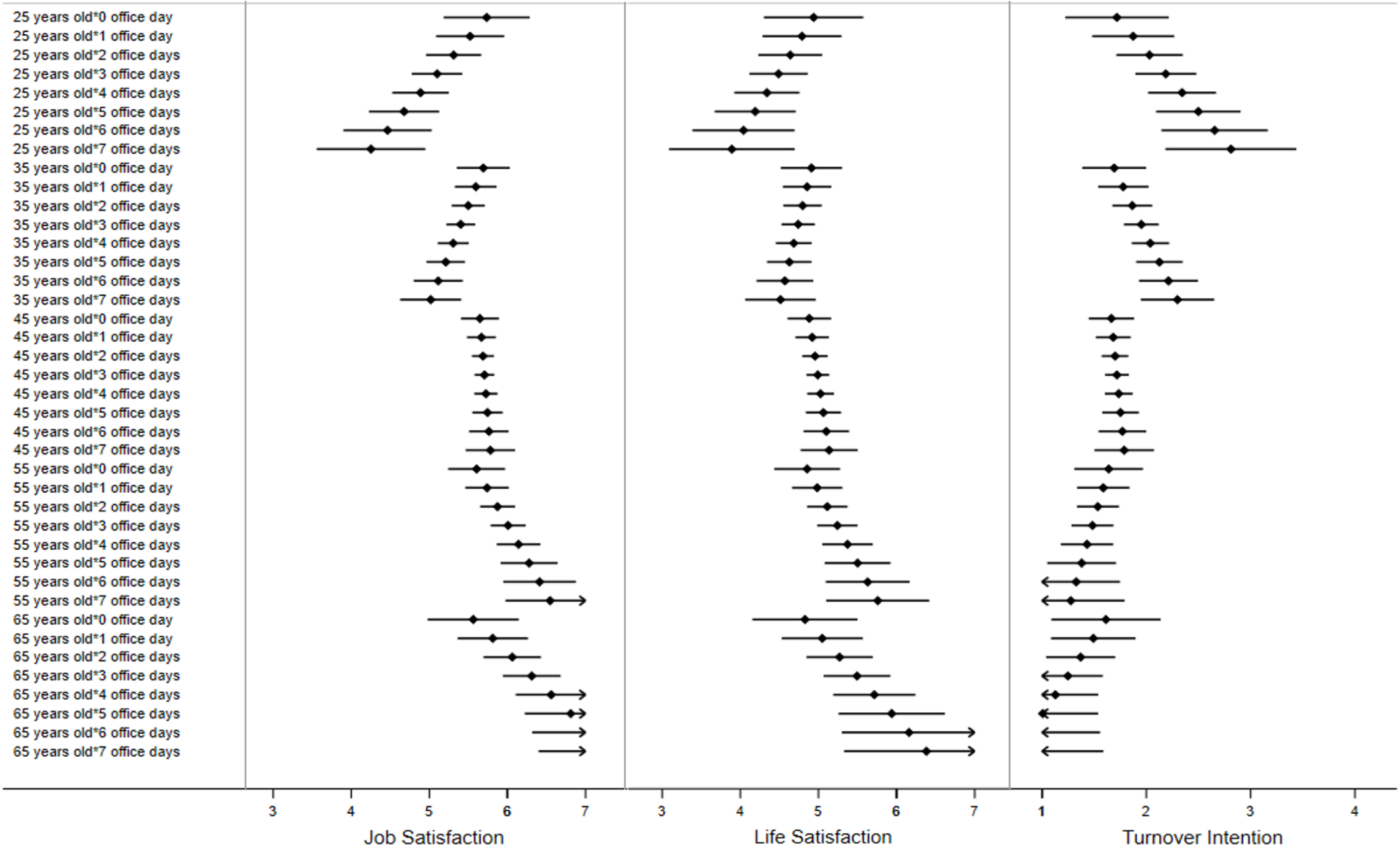
The predicted value and 95% confidence intervals (CIs) of job satisfaction, life satisfaction, and turnover intention by healthcare workers’ age and office days.

### 3.2 Predictors of job satisfaction, life satisfaction, and turnover intention

First, healthcare workers’ office days predicted their job satisfaction but the relationship depended on their age (b=0.01; 95% CI: 0.00 to 0.02; p=0.004). Margin analysis showed that office days negatively predicted job satisfaction for the young healthcare workers (e.g. at 25 years old: b=-0.21; 95% CI: −0.36 to −0.60; p=0.006). On the contrary, office days positively predicted job satisfaction for the older workers (e.g. at 65 years old: b=0.25; 95% CI: 0.06 to 0.44; p=0.009).

Second, office days predicted workers’ life satisfaction overall, and the relationship also depended on their age (b=0.01; 95% CI: 0.00 to 0.02; p=0.041). In particular, the number of office days was positively associated with life satisfaction among the older workers (e.g. at 65 years old: b=0.22; 95% CI: 0.01 to 0.44; p=0.043) but was marginally negatively associated with life satisfaction among the younger workers (e.g. at 25 years old: b=-0.15; 95% CI: −0.32 to 0.03; p=0.094). In addition, job redeployment (b=-0.51; 95% CI: −0.97 to −0.05; p=0.031) and daily exercise hours in the past week (b=0.07; 95% CI: 0.00 to 0.14; p=0.037) both predicted life satisfaction.

Lastly, healthcare workers’ office days predicted their turnover intention, and the relationship similarly depended on their age (b=-0.01; 95% CI: −0.01 to 0.00; p=0.05). In particular, the number of office days was positively associated with turnover intention among the younger workers (e.g. at 25 years old: b=0.16; 95% CI: 0.02 to 0.29; p=0.025) but not the older healthcare workers.

## 4. Discussion

Healthcare workers are especially critical in the COVID-19 pandemic. This study presents the first attempt to identify which healthcare workers have more or less job satisfaction, life satisfaction, and turnover intention during the COVID-19 pandemic.

Several covariates, which were significant predictors of mental health and well-being in healthcare workers in earlier studies, were not significant in our study. For instance, age and gender predicted mental health and well-being among healthcare workers in China (Lai *et al*., 2020, Li *et al*., in press, Liang *et al*., 2020) and in Iran (Kaveh *et al*., 2020), and in Spain (age only) (Romero *et al*., in press) during the COVID-19 pandemic, however neither age or gender predicted outcomes in this study. In addition, education was found to be a predictor of mental health of health staff in studies in China (Li *et al*., in press, Liu *et al*., in press) but not in another study in Iran (Zhang *et al*., in press). Education was not a significant predictor of satisfaction or turnover intention in our sample. These findings support that the predictors of healthcare staff’s job related variables and well-being during a pandemic vary across countries (Zhang *et al*., in press).

Moreover, this study identified several unique risk factors including weekly office days. Importantly, the number of office days predicted job satisfaction positively for the older healthcare staff, but negatively for the younger staff. Similar patterns existed on the outcome variables of life satisfaction, and turnover intention. These findings suggest healthcare organizations need to pay attention to younger workers working many days a week and older workers with fewer office days.

While it can be necessary to redeploy workers or hire temporary staff to deal with the COVID-19 crisis, those workers who were redeployed were less satisfied with their lives. Hence, hospitals may need to better support those redeployed healthcare workers. It is also worth noting the temporary healthcare staff did not significantly differ from regular staff on job satisfaction, life satisfaction, and turnover intention.

This study has some limitations. First, as data collection is challenging during the COVID-19 crisis, particularly for busy healthcare staff, we used convenience sampling. Second, to our best knowledge, this is the first study that has examined several job characteristics, such as office days, temporary staff, and job redeployment, as predictors for healthcare workers’ job-related outcomes under COVID-19. Such practices differ across countries and healthcare systems (Jahanshahi et al., 2020), and future studies may test these factors.

Protecting and retaining healthcare workers is paramount during a pandemic. This study demonstrates healthcare workers’ number of office days matters to their job satisfaction, life satisfaction, and turnover intention. However, the number of office days carries different impacts for younger and older workers. We call for more studies on healthcare workers from the perspective of their ongoing working characteristics under the COVID-19 pandemic.

## Data Availability

available upon request.

## Declaration of Competing Interest

The authors declare that there are no potential conflicts of interest with respect to the research, authorship, and/or publication of this article.

## Acknowledgement

We acknowledge the support of Tsinghua University-INDITEX Sustainable Development

Fund (Project No. TISD201904).

## Notes

### Competing Interest Statement

The authors have declared no competing interest.

### Clinical Trial

This study has not involved of all clinical trials.

### Author Declarations

The study was approved by the ethics committee of Tsinghua University (20200322).

